# A Novel Intervention Recurrent autoencoder for real time forecasting and non-pharmaceutical intervention selection to curb the spread of Covid-19 in the world

**DOI:** 10.1101/2020.05.05.20091827

**Authors:** Qiyang Ge, Zixin Hu, Shudi Li, Wei Lin, Li Jin, Momiao Xiong

**Author notes:** Address for correspondence and reprints: Dr. Momiao Xiong, Department of Biostatistics and Data Science, School of Public Health, The University of Texas Health Science Center at Houston, P.O. Box 20186, Houston, Texas 77225.

## Abstract

As the Covid-19 pandemic soars around the world, there is urgent need to forecast the number of cases worldwide at its peak, the length of the pandemic before receding and implement public health interventions to significantly stop the spread of Covid-19. Widely used statistical and computer methods for modeling and forecasting the trajectory of Covid-19 are epidemiological models. Although these epidemiological models are useful for estimating the dynamics of transmission od epidemics, their prediction accuracies are quite low. To overcome this limitation, we formulated the real-time forecasting and evaluating multiple public health intervention problem into forecasting treatment response problem and developed recurrent neural network (RNN) for modeling the transmission dynamics of the epidemics and Counterfactual-RNN (CRNN) for evaluating and exploring public health intervention strategies to slow down the spread of Covid-19 worldwide. We applied the developed methods to the real data collected from January 22, 2020 to May 8, 2020 for real-time forecasting the confirmed cases of Covid-19 across the world.

## Introduction

As of May 11, 2020, global confirmed cases of Covid-19 passed 4,152,670 and has spread to 212 countries, causing fear globally (Anastassopoulou et al. 2020). The serious public health threat of Covid-19 has never been seen for more than one century. The government officers and people around the world are desperately trying to slow the spread of Covid-19 (Irfan 2020). We must change our policies to deal with increased mobility of citizens and immediately implement the public health interventions and expanding the virus testing to stop the spread of Covid-19 across the world. How computer modeling of Covid-19’s transmission dynamics could help governments to quickly and strongly move to slow down the spread of Covid-19?

Widely used statistical and computer methods for modeling of Covid-19 simulate the transmission dynamics of epidemics to understand their underlying mechanisms, forecast the trajectory of epidemics, and assess the potential impact of a number of public health measures on curbing the spread speed of Covid-19 (Li et al. 2020, Wu et al. 2020, Zhao et al. 2020, Kucharski et al. 2020, Tuite et al. 2020, Hellewell et al. 2020, Li et al. 2020). Although these epidemiological models are useful for estimating the dynamics of transmission, and evaluating the impact of intervention strategies, they have some serious limitations (Funk et al. 2018, Johansson et al. 2019). First, the epidemiological models consist of ordinary differential equations that have many unknown parameters, and depend on many assumptions. It is difficult to translate public interventions to these parameters. Most analyses used hypothesized parameters, which often lead to fitting data very poor. Health officers desperately want to track the trajectory of epidemics and accurately estimate the peak time and number of cases, duration, and ending time and number of cases of Covid-19 for their health policy plan. However, the forecasting results of using the classical epidemiological models such as Susceptible-Exposed-Infectious-Removed (SEIR) models are highly unreliable. Second, the successful application of public health intervention planning highly depends on the model parameter identifiability. However, overall, the parameters in the complex compartmental dynamic models are unidentifiable (Roosa and Chowell 2019, Roda et al. 2020). The values of parameters cannot be uniquely determined from the real data (Gábor et al. 2017). The variances of the estimators of these parameters are very high.

To overcome limitations of the epidemiological model approach, and assist public health planning and policy making, we formulated the real-time forecasting and evaluating multiple public health intervention problem into off-policy evaluation (OPE) and forecasting treatment response problem where the aim is to estimate the response of a new public health intervention policy, given historical data that may have been generated by a different public health intervention policies (Bibaut et al. 2019). We viewed the interventions as treatments where multiple interventions were administered at different time points. The number of new cases were taken as treatment responses. The ability to accurately estimate effects of public health interventions over time would allow health officers to determine what intervention strategies should be used and the optimal time at which to implement them (Lim et al. 2018). Recurrent Intervention Network (RIN) (Lim et al. 2018) where a recurrent neural network architecture for forecasting a nation’s response (number of new cases) to a sequence of planned interventions were used to forecast and evaluate multiple public health interventions for Covid-19 worldwide. Potential outcomes of RIN were trajectory of the spread of Covid-19. Public health interventions including locking down residential buildings and compounds, strict self-quarantine for families, door-to-door inspection for suspected cases, maintaining social distancing, stopping mass gatherings, closure of schools and universities, vacating hotels and university dormitories. To quantify comprehensive intervention strategies, an intervention variable that comprehensively and abstractly measures mobility activities and social distancing was used as an input variable for each block of RIN (detailed description of the intervention measure was summarized in Training Procedures and Loss function Section in the Supplementary Note A). We cluster all the countries in the world into several groups. For each group, a value (weight) was assigned to each group such that the average prediction error of counterfactual recurrent network (CRN) was small. The RIN is taken as a general framework for investigating how Covid-19 evolves under different intervention plans, how individual nation responds to intervention over time, but also which are optimal timings for assigning interventions. Therefore, this approach will provide new tools to improve public health planning and policy making.

The RIN was applied to the surveillance data of lab confirmed Covid-19 cases in the world up to May 8, 2020. Data on the number of confirmed, new and death cases of Covid-19 from January 22, 2020 to May 8, 2020 were obtained from John Hopkins Coronavirus Resource Center (https://coronavirus.jhu.edu/MAP.HTML).

## Methods

### RIN as a Framework for modeling and forecasting the spread of Covid-19 over time with multiple interventions

The RIN uses sequence-to-sequence multi-input/output recurrent neural network (RNN) architectures to model health intervention plan and make multi-step prediction of the response trajectory of Covid-19 over time with multiple interventions (Lim et al., 2018). The RNN can learn the complex dynamics within the temporal ordering of input time series of Covid-19 and use an internal memory to remember. The health intervention plan has multiple intervention regimens. As shown in Figure 1, the RIN determines the intervention response (similar to counterfactual outputs) for a given set of planned interventions and evaluates the impact of different intervention strategies and their implementation times on the curbing the spread of Covid-19 and provides timely selection of optimal sequence of intervention strategies.

**Figure 1.**
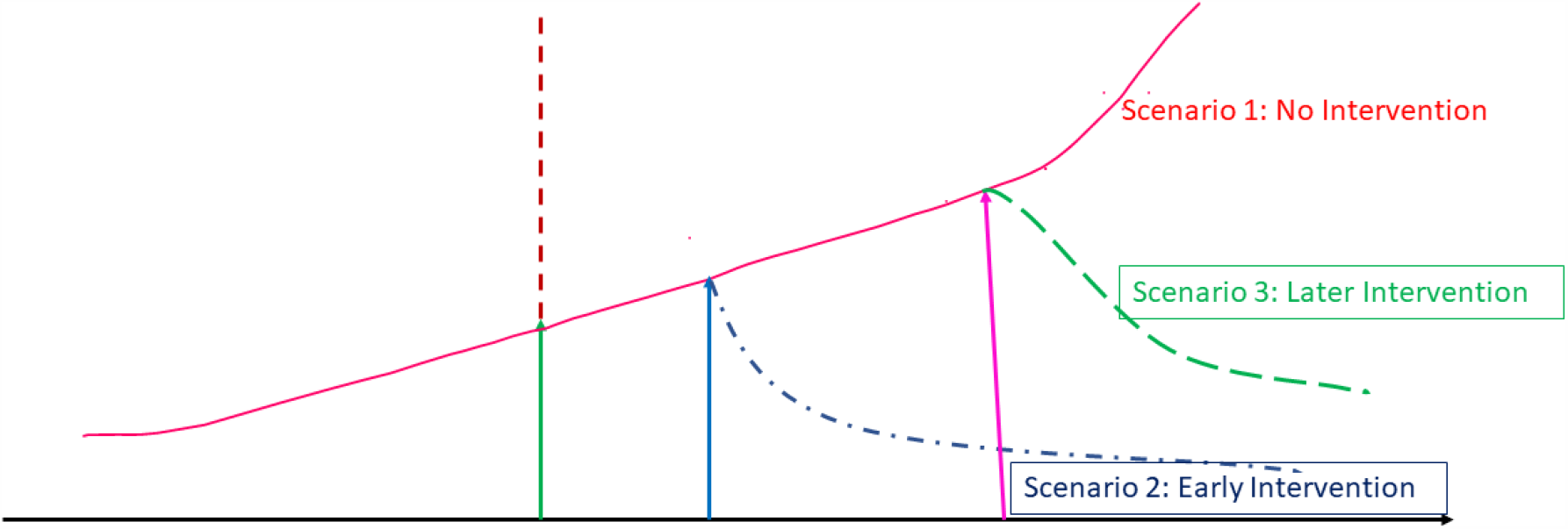
Forecasting intervention response to curbing the spread of Covid-19 under a sequence of interventions.

The RIN is a RNN autoencoder. It consists of two RNNs: the encoder RNN (vanilla RNN (Figure S1) or long short-term memory (LSTM) (Figure S2) is used as encoder) and the decoder RNN (vanilla RNN or LSTM is used as the decoder). The RNN encoder models input time series (past history of the number of cases of Covid-19 over time) and predicts future response time series (number of cases of Covid-19 in the future with a planned sequence of interventions) (Srivastava et al., 2015). The latent state of the RNN encoder after reading in the entire input time series (past trajectory of Covid-19), is the representation (compressed latent features of the entire input time series) of the input trajectory of Covid-19. Unlike the standard decoder where the decoder reconstructs back the input time series from the latent representation, the RNN decoder uses the learned features of the dynamics of Covid-19 in the RNN encoder to forecast the potential response time series, given a sequence of planned public health interventions as an input to the RNN decoder. The feature vector learned in the RNN encoder is then provided as an input to the RNN decoder which initiate prediction of the future dynamics of Covid-19 under the future interventions (Figure 2). Detailed description of the RIN was listed in Supplementary Note A.

**Figure 2.**
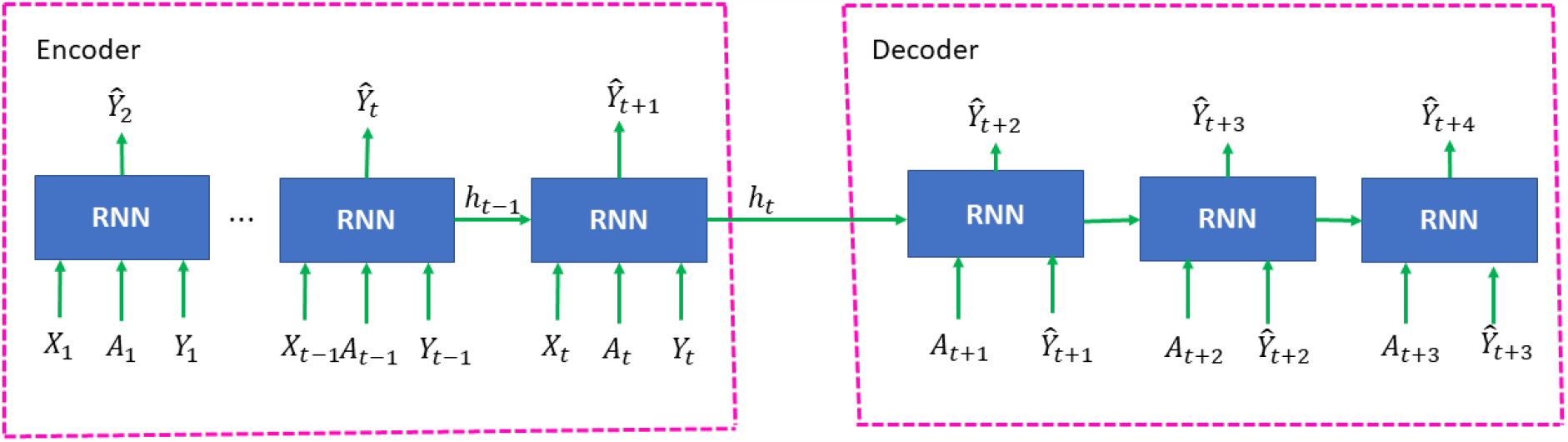
Architecture of recurrent intervention network.

### Training the RIN

RIN training consisted of RNN encoder training and RNN decoder training (Supplementary Note A). We first fitted a model of the system’s dynamics of Covid-19 to the data from past experimental interventions to learn representations of the states of the dynamics of Covid-19 (encoder training), and then used the learned fit to extrapolate and forecast the response to the alternative interventions (decoder training).

The RNN encoder training procedures were briefly introduced here. For details, please see the Supplementary Note A. The basic RNN unit in the RIN consisted of input layer, hidden layer and output layer (Figure S1). The input variables in the RNN encoder can include covariates *X*_*t*_ such as density of population, traffic flow, health facility resources, GDP, and social-economic status although this study did not include these quantities, intervention variable *A*_*t*_ and the numbers of cases (potential outcomes) *Y*_*t*_ at the time *t*. The state in the hidden layer at the time *t* was denoted by *h*_*t*_. The output layer had the output variable *Y*_*t*+1_. A nonlinear activation function was exponential linear unit (ELU) (Clevert et al., 2015) which was defined as

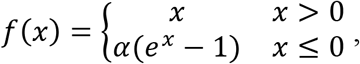

where *α* > 0.

ELU is similar to ReLU when *x* > 0. ELUs diminish the vanishing gradient effect as ReLUs. The vanishing gradient problem is alleviated because the positive part of these functions is the identity, therefore their derivative is one and not contractive. However, tanh and sigmoid activation functions are contractive almost everywhere.

In contrast to ReLUs, ELUs have negative values (ReLU does not have), which pushes the mean of the activations closer to zero. Mean activations that are closer to zero enable faster learning as they bring the gradient closer to the natural gradient.

The input data were divided into several batches with length of 7 days. Each batch was used to train the RNN encoder which forecasted standard one-step-ahead intervention response *Ŷ*_*t*+1_ as close to the observed intervention response *Y*_*t*+1_ as possible via the nonlinear mapping (Supplementary note A)

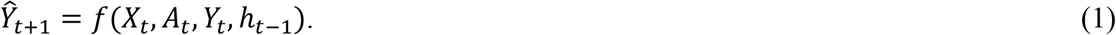

The mean-squared error was used as loss function for training the RNN encoder. The training was performed via the standard propagation algorithms (Supplementary A). After training was completed, the RNN encoder extracted the hidden state *h*_*t*_that captured the internal features of the transmission dynamics of Covid-19 via performing a feed-forward pass over the training data on the RNN encoder (Supplementary A).

After the RNN encoder training was completed, we began to train the RNN decoder. An RNN unit in the RNN decoder consisted of input layer with intervention variable *A*_*t*+*τ*_, (*τ* = 1,2, …), hidden layer with hidden state *Z*_*t*+*τ*−1_ and output *Y*_*t*+*τ*+1_. For a given country, observations, intervention *A*_*t*_ and the number of cases *Y*_*t*_ were randomly divided into short batches of up to *τ*_*b*_ time steps. Each batch of short sequence starting at time *t* and ending at time *t* + *τ*_*b*_ − 1 consisted of 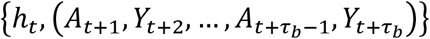. The mean square errors were still used as the RNN decoder loss function. The goal of RNN decoder training is to make its training loss function smallest (Supplementary A).

### Forecasting Procedures

After completion of the training, the trained RIN was used to forecast the future number of cumulative cases of Covid-19 under some planned interventions for each country. During evaluation, we do not have access to ground-truth outcomes. Therefore, we used the trained decoder to make one step ahead forecasting. The outcomes forecasted by the decoder (*Ŷ*_*t*+1_, …,*Ŷ*_*t*+*τ*−1_) were recursively used as inputs. The recursive multiple-step forecasting involved using a one-step model multiple times where the prediction for the preceding time step and intervention strategy were used as an input for making a prediction on the following time step. For example, for forecasting the number of new confirmed cases for the one more next day, the predicted number of new cases in one-step forecasting would be used as an observational input in order to predict day 2. Repeat the above process to obtain the two-step forecasting. The summation of the final forecasted number of new confirmed cases for each country was taken as the prediction of the total number of new confirmed cases of Covid-19 worldwide under the intended intervention. By running the decoder, we can select starting and ending time of different interventions and the optimal or appropriate interventions to give over time to obtain the best outcomes of controlling the spread of Covid-19 for each country.

### Data Collection

The analysis is based on surveillance data of confirmed cumulative and new COVID-19 cases worldwide as of May 8, 2020. Data on the number of cumulative and new cases and COVID-19-attributed deaths across 186 countries from January 22, 2020 to May 8, 2020 were obtained from John Hopkins Coronavirus Resource Center (https://coronavirus.jhu.edu/MAP.HTML).

### Data Pre-processing

log_2_ was used to transform the original data: 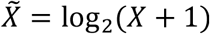. The intervention measure was calculated as follows. Set the intervention measure at the final time 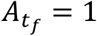 for China, 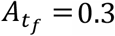 for Korea South, Switzerland, United Kingdom, Spain, US, Italy, Germany, Iran, and France, and 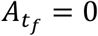 for all other countries. Assume that the intervention measure curve is an exponential function starting at 0 and ends at 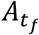. The intervention measure *A*_*t*_ is given by 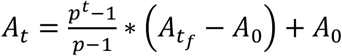, where *p* > 0, *p* ≠ 1 is the curve shape factor and *t* takes values in evenly sliced numbers of interval [0, 1], *A*_0_ is the intervention measure at the initial time *t*_0_. When *p* = 1, *A*_*t*_ is a linear function 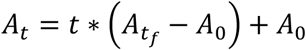. In this study, we set *p* = 0.01.

We randomly picked *k* = 64 countries with *l* = 7 length of Covid-19 time series (the number of new cases over time) data staring from the same day to generate *k* time series with *l* length for a minibatch to be used for backpropagation training through time. Calculate the mean value of each time series in the batch. The values of each time series were divided by their mean values to normalize the data.

## Results

### Prediction accuracy of dynamics of Covid-19 using RIN

Accurate prediction of the spread of Covid-19 is important for health intervention plan for the future. To demonstrate that the RIN is an accurate forecasting method, the RIN was applied to confirmed accumulated cases of COVID-19 across 186 countries. Figure 3 plotted reported and one-step ahead predicted time-case curves of Covid-19 where blue dotted curve was the number of estimated cumulative cases after the analysis completion. To further reliably evaluate the forecasting accuracy, we reported 10-step ahead forecasted numbers of cumulative cases and errors of Covid-19 of 8 countries in Table 1 starting with April 29, 2020. The forecasting errors were quite small.

**Table 1.**
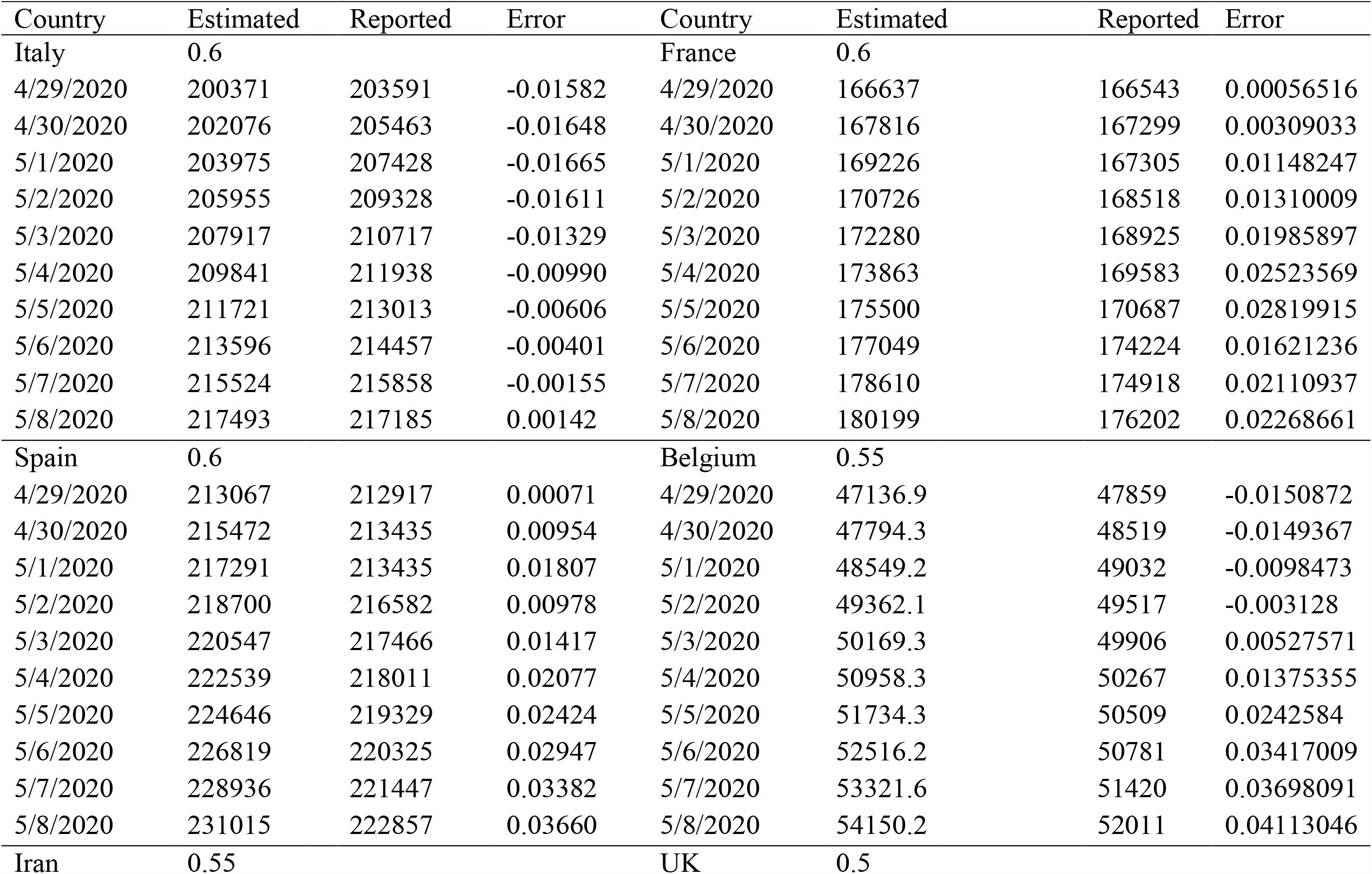
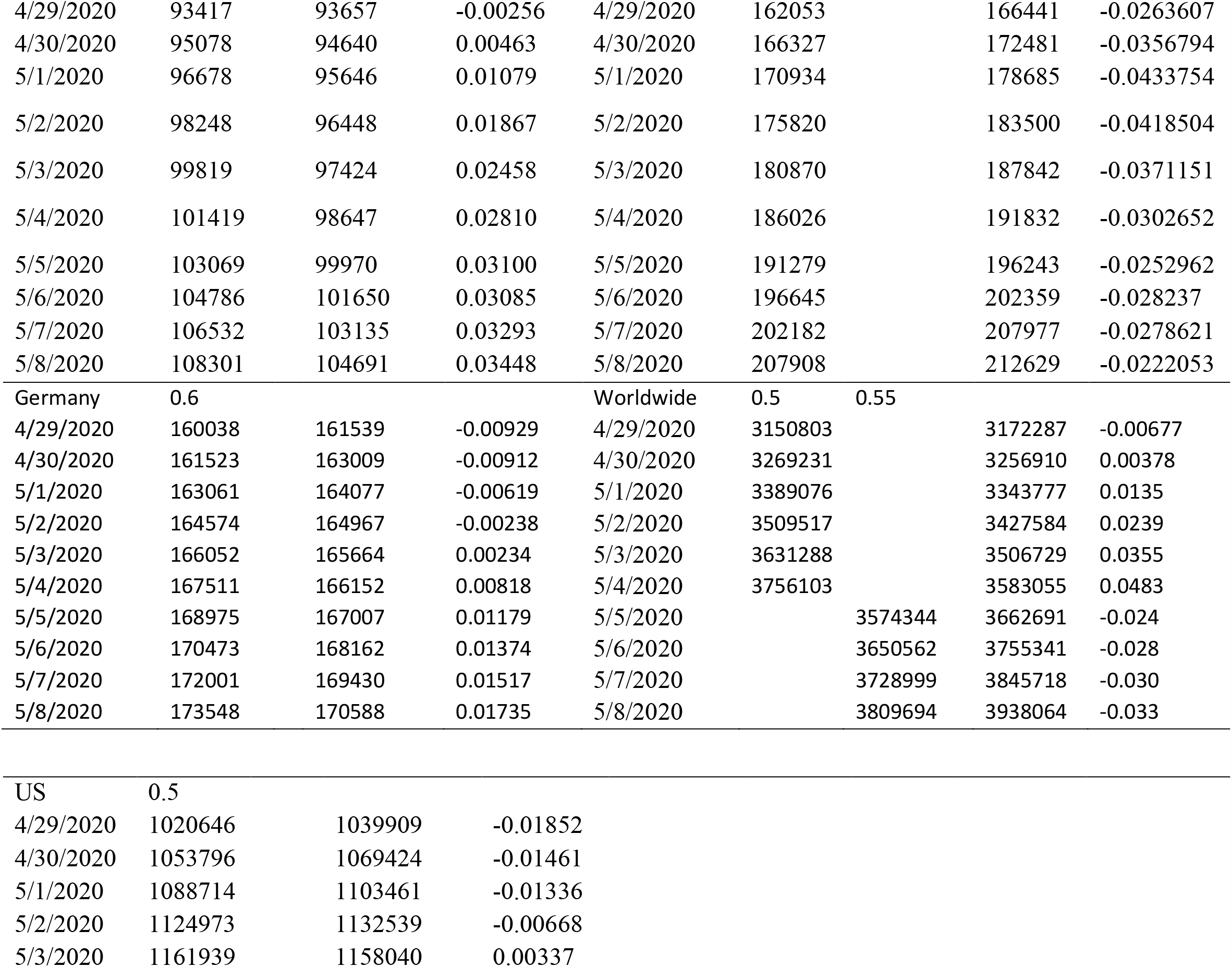
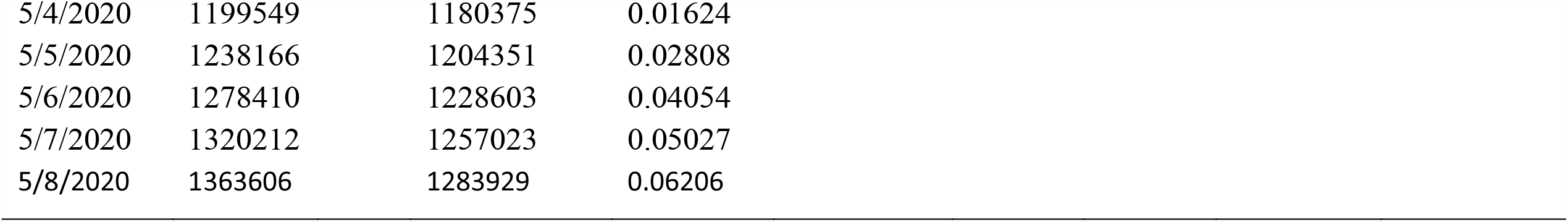
Forecasting errors of worldwide and eight countries, where 0.5, 0.55 and 0.6 were intervention measures

**Figure 3.**
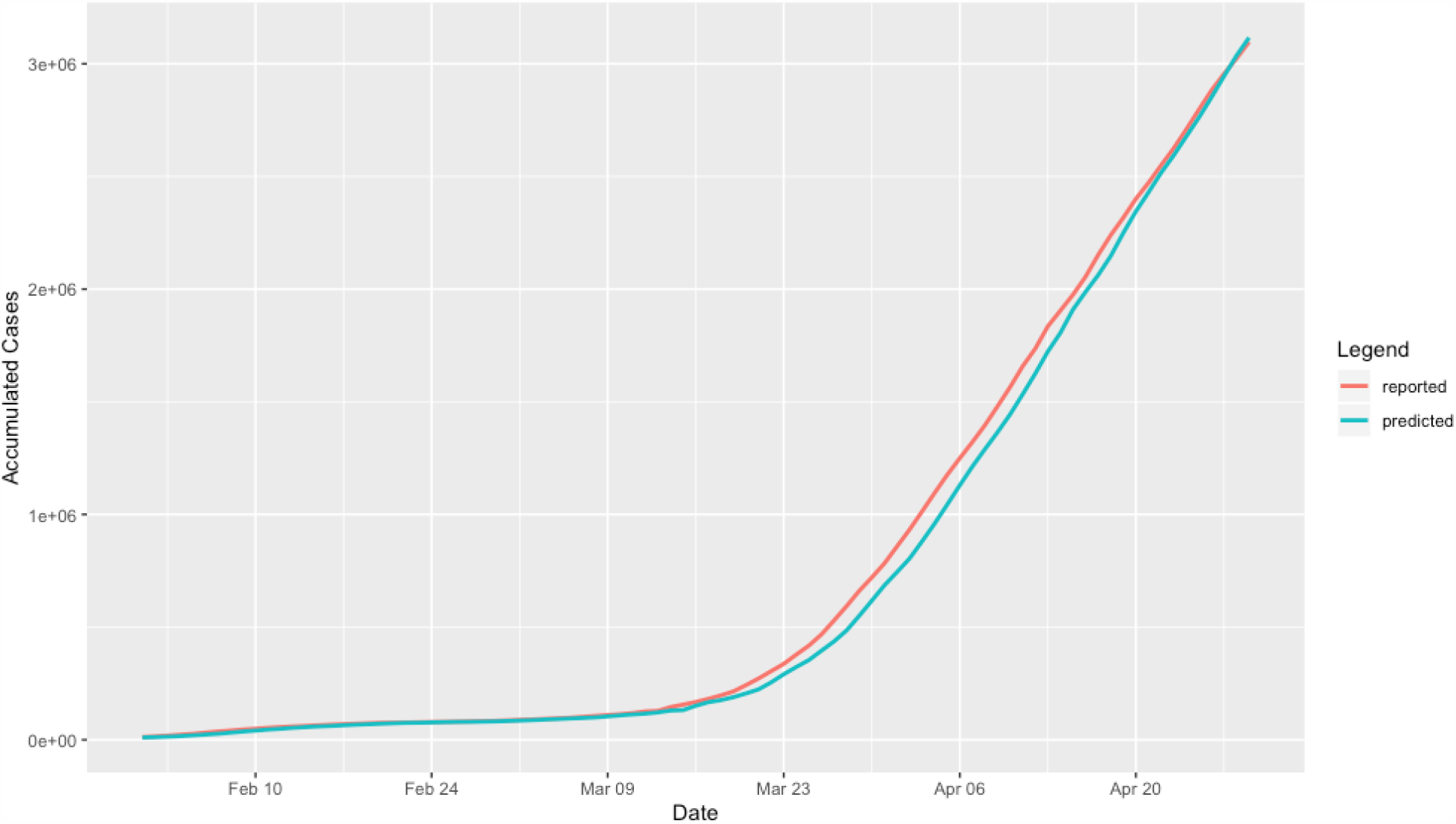
Reported and predicted time-case curves of Covid-19 worldwide where blue dotted curve was the number of reported cumulative cases after the analysis completion.

Inputting a sequence of hypothesized public health intervention strategies, the outputs of the RIN decoder were counterfactual numbers of cases of Covid-19 to respond to the intervention strategies. Interventions were measured by number in the interval [0, 1], where 1 indicated the strictest comprehensive public health intervention, 0 indicated no intervention and the values between 0 and 1 indicated the various less strict interventions. To intuitively illustrate the impact of the measure of intervention on the spread of Covid-19, we presented Figure S3. Figure S3 plotted counterfactual numbers of new cases of Covid-19 over time worldwide to respond the interventions with values 0, 0.3, 0.5, 0.7 and 1. We observed that if the measure of intervention was 1, the number of new cases was dramatically deceased to zero. However, when the measure of intervention was 0.3, the number of new cases exponentially increased. The measure of intervention had big effect on the spread of Covid-19.

The number of cases of Coid-19 was a function of the past history and the measure of intervention. Forecasting also depended on the measure of intervention. In Table 1, we also listed the measures of the interventions which provided information on the degrees of current interventions in the country. The measure of interventions in the most countries was 0.6. However, the current measure of interventions in UK was 0.5, the smallest in 8 countries. These results showed that the RIN for forecasting the trajectory of Covid-19 was accurate and reliable. Similar to causal inference, the RIN can be used to evaluate the impact of a sequence of multiple intervention strategies on the curbing the spread of Covid-19 if the interventions were viewed as treatments.

### The number of cases of Covid-19 grows exponentially without additional intensive interventions

To investigate how Covid-19 pandemic surges around the world, we presented Figure 4 that showed the forecasted number of cumulative cases of Covid-19 worldwide over time, assuming that the current intervention measure remains. We observed that the number of cumulative cases of Covid-19 exponentially grown and would reach extremely high number 8,491,301 on July 1, 2020 if none of additional comprehensive public health intervention was implemented. Similarly, Figure 5 and Figure S3 plotted time-case curves of Covid-19 of eight countries: Italy, Spain, Iran, Germany, USA, France, Belgium and UK, and worldwide with unchanged intervention strategies in the future, respectively. We also observed exponentially growth of the numbers of cases of Coid-19 for many countries without additional intervention.

**Figure 4.**
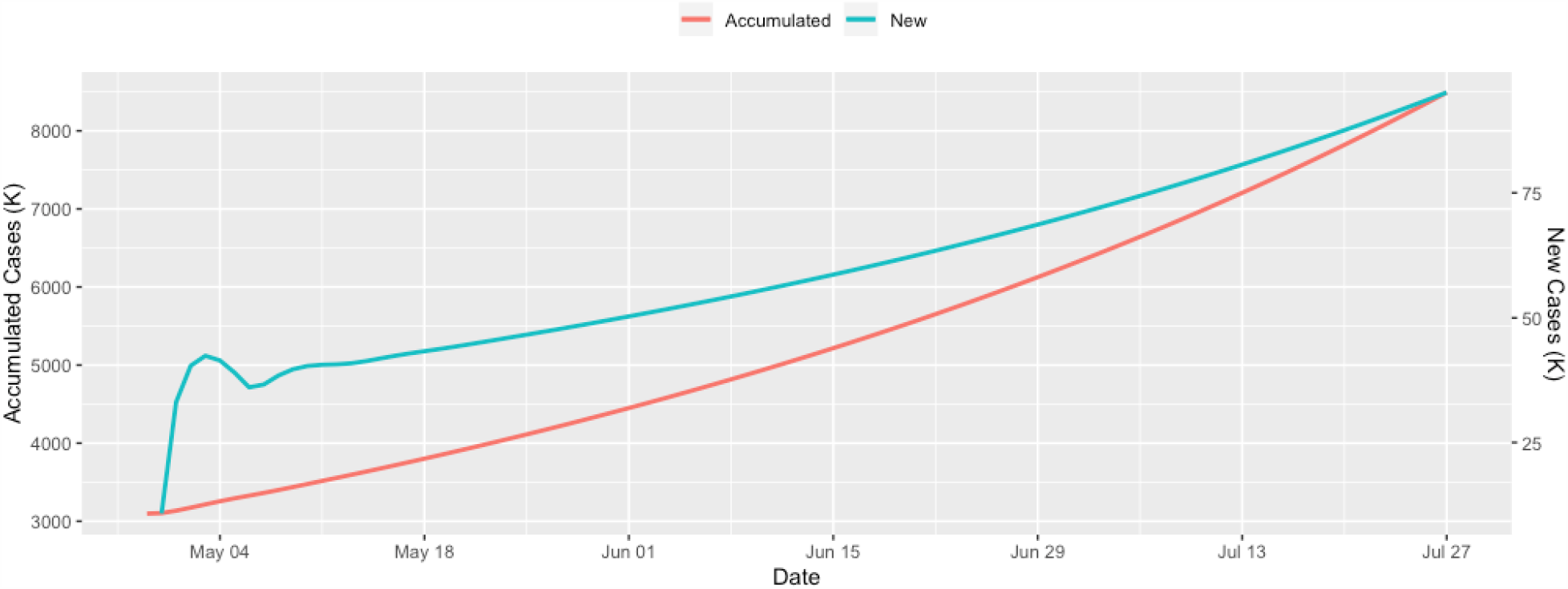
The numbers of cumulative and new cases of Covid-19 worldwide over time, assuming the current intention remains unchanged. The curves in blue color and red color represented the number of cumulative cases and the number of new cases, respectively.

**Figure 5.**
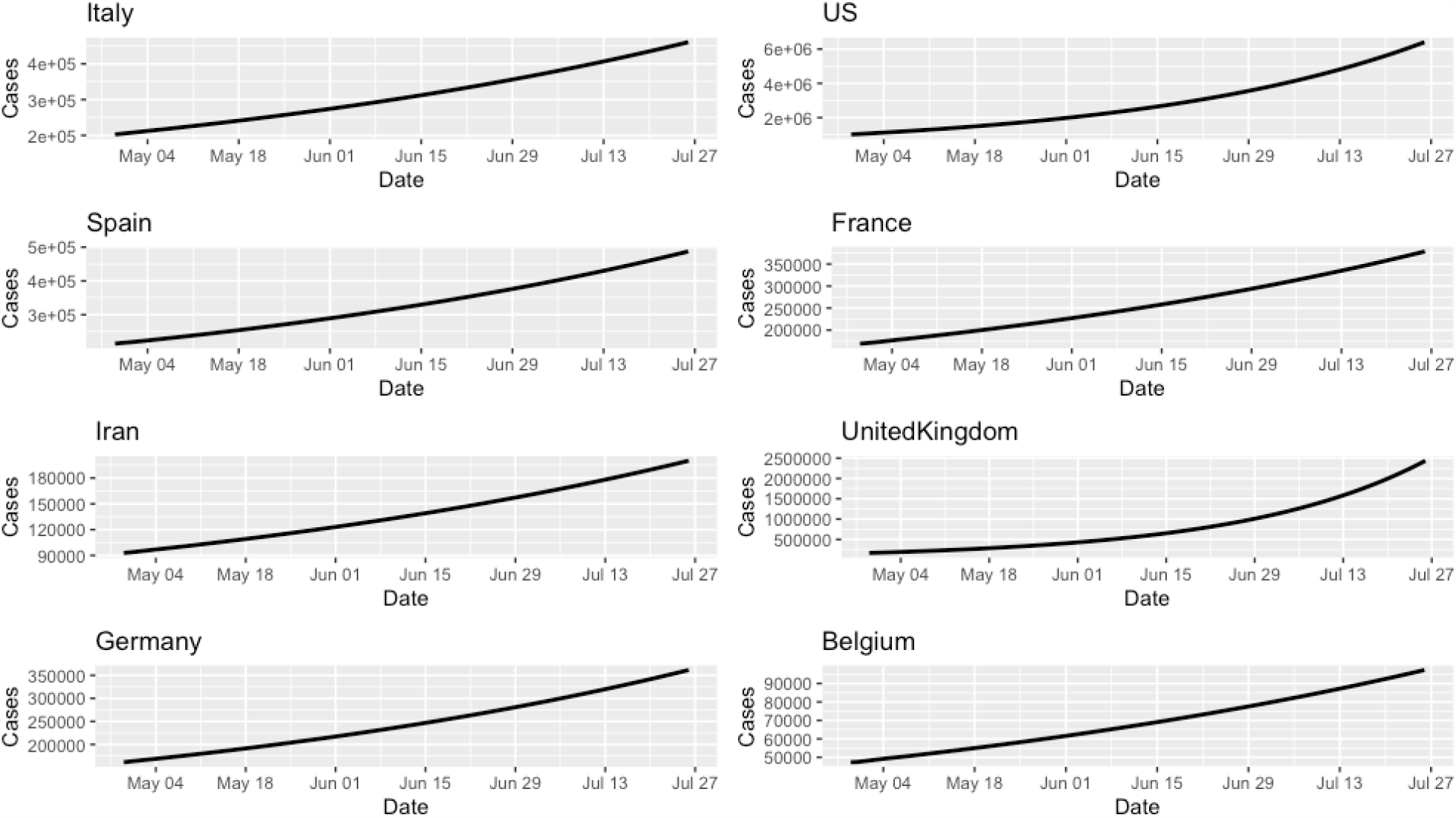
Forecasted number of cumulative cases of Covid-19 of eight countries Italy (A), Spain (B), Iran (C), Germany (D), USA (E), France (F), UK (G) and Belgium (H) over time without additional interventions.

### We are in the eve to successfully curb the spread of Covid-19

As Covid-19 Accelerates and exponentially grows, how to slow down the spread of Covid-19 is an urgent task for every country around world. To demonstrate that when the additional intervention was implemented, the number of new cases of Covid-19 would decrease, we presented Figures 6 and 7. Figures 6 and 7 plotted the number of cumulative case and new case curves of Covid-19 over time for 12 countries: US, Italy, Spain, Germany, France, Iran, UK, Switzerland, Belgium, South Korea, Japan and Singapore under three invention scenarios, respectively. Scenario 1 started with the intervention measure of 0.50 for one week, then transitioned to the intervention with a measure of 0.70 in two weeks. The scenario 2 started with the intervention measure of 0.5 for the first week, and then transitioned to the intervention with a measure of 0.70 in three weeks. The scenario 3 started with the intervention measure for one week, changed to 0.60 in three weeks and finally transitioned to the intervention with a measure of 0.70 in three weeks. Figures 6 and 7 showed that when the countries moved to intervention with measure of 0.70, the spread of Covid-19 in all 12 countries was curbed. Now the measures of interventions in the most of 12 countries were closer to 0.60 (Table 1). These countries were closing to stopping the spread of Covid-19 if additional interventions such as wearing face masks were implemented.

**Figure 6.**
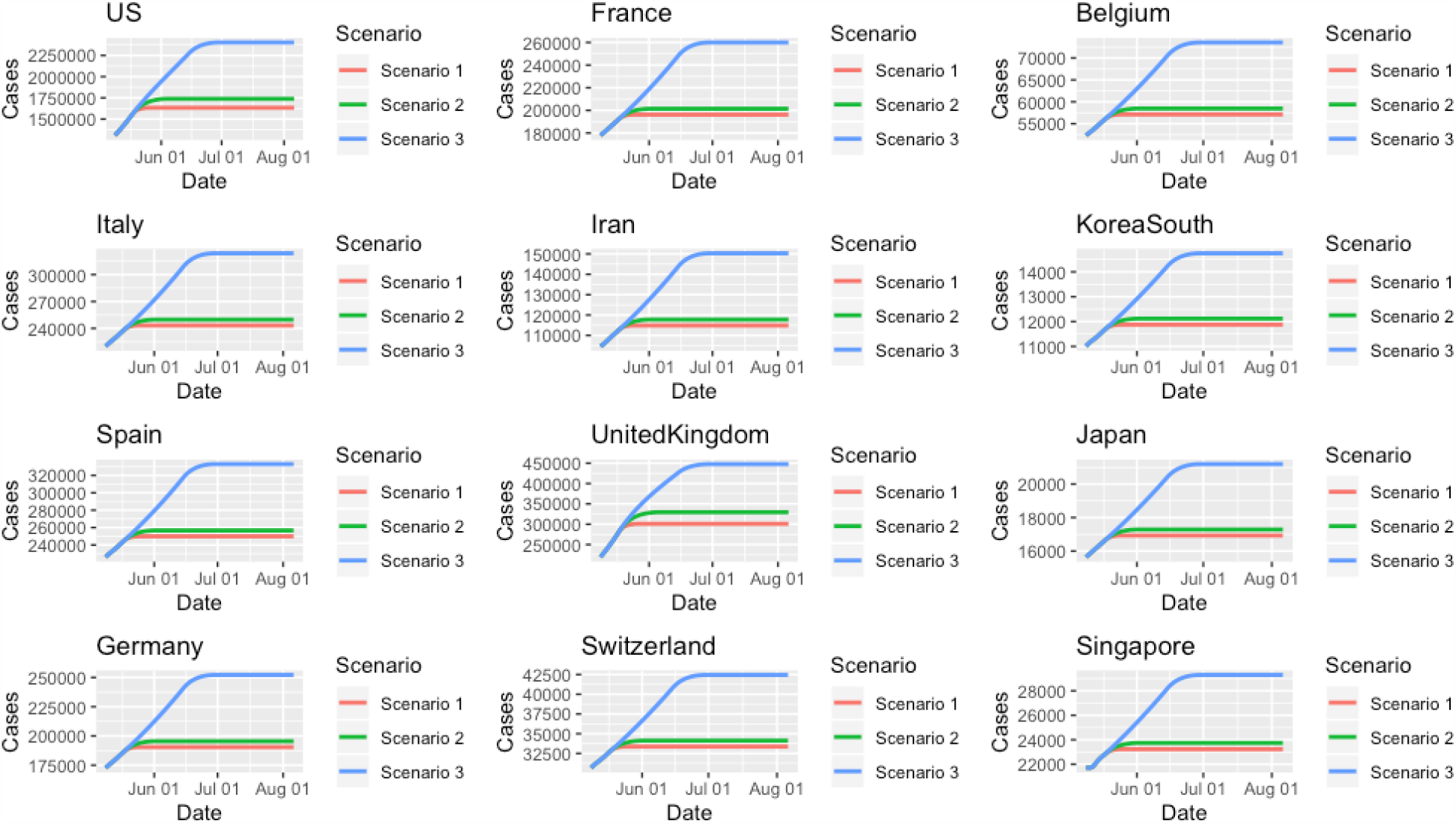
Number of cumulative case curves of Covid-19 over time for 12 countries under three invention scenarios. (A) Time-case plot for US, (B) Time-case plot for Italy, (C) Time-case plot for Spain, (D) Time-case plot for Germany, (E) Time-case plot for France, (F) Time-case plot for Iran, (G) Time-case plot for UK, (H) Time-case plot for Switzerland, (I) Time-case plot for Belgium, (J) Time-case plot for South Korea, (K) Time-case plot for Japan, and (L) Time-case plot for Singapore.

**Figure 7.**
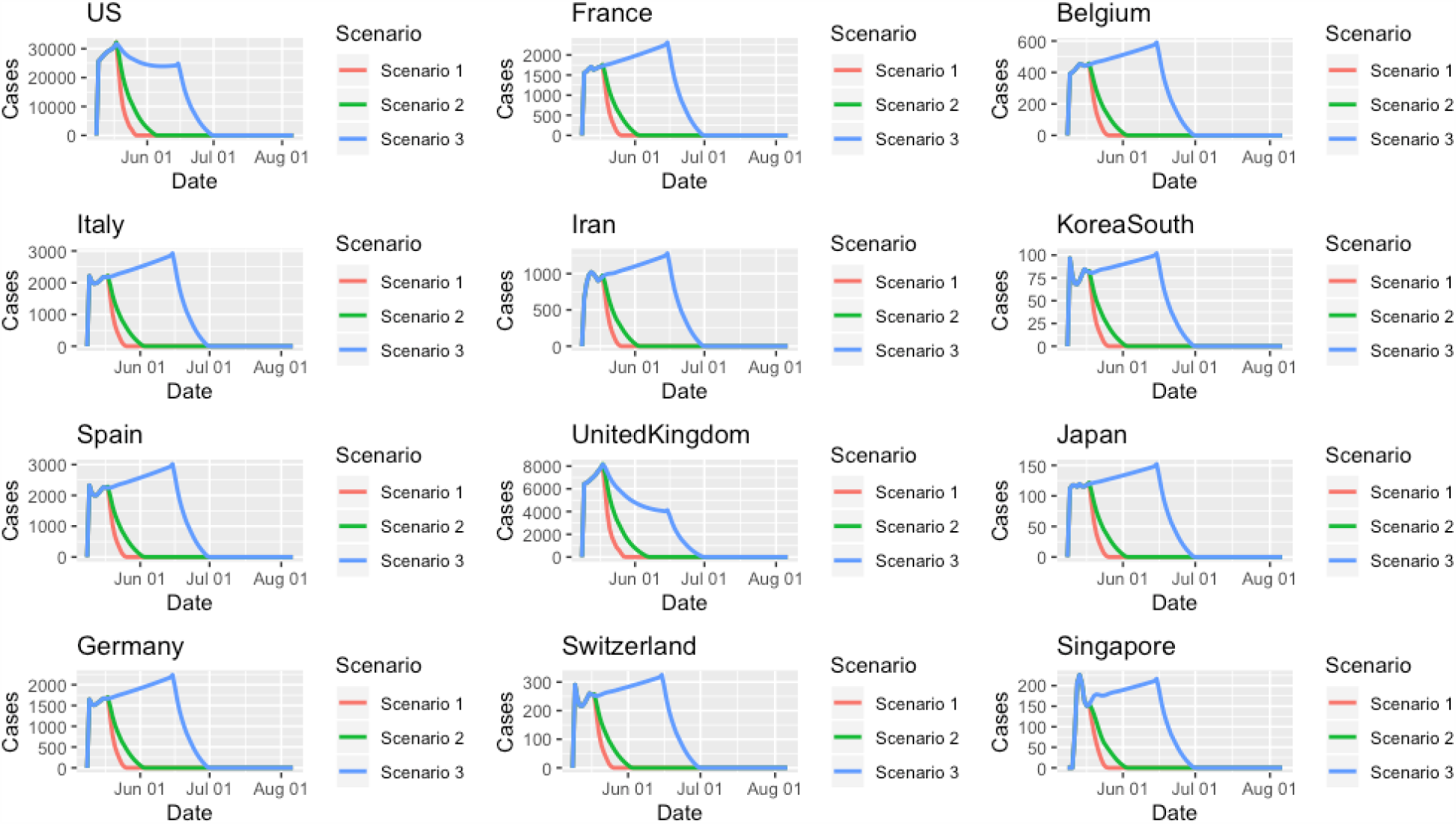
Number of new case curves of Covid-19 over time for 12 countries under three invention scenarios. (A) Time-new case plot for US, (B) Time-new case plot for Italy, (C) Time-new case plot for Spain, (D) Time-new case plot for Germany, (E) Time-new case plot for France, (F) Time-new case plot for Iran, (G) Time-new case plot for UK, (H) Time-new case plot for Switzerland, (I) Time-new case plot for Belgium, (J) Time-new case plot for South Korea, (K) Time-new case plot for Japan, and (L) Time-new case plot for Singapore.

Next we investigated how various intervention strategies reduced the peak time and cumulative case numbers, and the final total number of cases. Table 2 showed the forecasted results of COVID-19 worldwide and in 11 countries under three sequences of interventions (Scenarios 1-3). Figure S4 plotted the time-case curves of Covid-19 worldwide under three invention scenarios. We can see that under all three scenarios, the peak times worldwide and in all 11 countries were May 18, 2020 and April 24 or before April 24, 2020, respectively; and the spread of COVID-19 worldwide and in all 11 countries would be stopped by the end of June, 2020.

**Table 2.** Covid-19 dynamics in 11 countries and worldwide for three scenarios.

| Scenario | Country | US | Italy | Spain | Germany | France | Iran |
| --- | --- | --- | --- | --- | --- | --- | --- |
| Scenario 1 | Peak Time | 4/24/2020 | 3/21/2020 | 3/25/2020 | 3/27/2020 | 4/12/2020 | 3/30/2020 |
|  | Peak Number (New) | 36188 | 6557 | 9630 | 6933 | 26849 | 3186 |
|  | Peak Number ( Cumulative) | 905358 | 53578 | 49515 | 50871 | 121712 | 41495 |
|  | End Time | 5/27/2020 | 5/26/2020 | 5/26/202 | 5/26/202 | 5/26/202 | 5/26/202 |
|  | End Cases | 1633438 | 243198 | 249919 | 190450 | 196231 | 114863 |
|  | Duration | 124 | 116 | 115 | 120 | 123 | 97 |
| Scenario 2 | Peak Time | 4/24/2020 | 3/21/2020 | 3/25/2020 | 3/27/2020 | 4/12/2020 | 3/30/2020 |
|  | Peak Number (New) | 36188 | 6557 | 9630 | 6933 | 26849 | 3186 |
|  | Peak Number ( Cumulative) | 905358 | 53578 | 49515 | 50871 | 121712 | 41495 |
|  | End Time | 6/6/2020 | 6/3/2020 | 6/3/2020 | 6/3/2020 | 6/3/2020 | 6/3/2020 |
|  | End Cases | 1738794 | 249776 | 256694 | 195499 | 201447 | 117782 |
|  | Duration | 134 | 124 | 123 | 128 | 131 | 105 |
| Scenario 3 | Peak Time | 4/24/2020 | 3/21/2020 | 3/25/2020 | 3/27/2020 | 4/12/2020 | 3/30/2020 |
|  | Peak Number (New) | 36188 | 6557 | 9630 | 6933 | 26849 | 3186 |
|  | Peak Number ( Cumulative) | 905358 | 53578 | 49515 | 50871 | 121712 | 41495 |
|  | End Time | 7/1/2020 | 7/1/2020 | 7/1/2020 | 7/1/2020 | 7/1/2020 | 7/1/2020 |
|  | End Cases | 2402819 | 323821 | 332980 | 252159 | 259993 | 150317 |
|  | Duration | 159 | 152 | 151 | 156 | 159 | 133 |
|  | Country | UK | Belgium | Korea South | Japan | Singapore | Worldwide |
| Scenario 1 | Peak Time | 4/10/2020 | 4/15/2020 | 3/3/2020 | 4/17/2020 | 4/20/2020 | 5/18/2020 |
|  | Peak Number (New) | 8733 | 2454 | 851 | 1161 | 1426 | 200938 |
|  | Peak Number ( Cumulative) | 74605 | 33573 | 5186 | 9787 | 8014 | 5685045 |
|  | End Time | 7/1/2020 | 7/1/2020 | 7/1/2020 | 7/1/2020 | 7/1/2020 | 5/28/2020 |
|  | End Cases | 300936 | 57109 | 11875 | 16927 | 23228 | 6116772 |
|  | Duration | 118 | 112 | 123 | 121 | 123 | 125 |
| Scenario 2 | Peak Time | 4/10/2020 | 4/15/2020 | 3/3/2020 | 4/17/2020 | 4/20/2020 | 5/18/2020 |
|  | Peak Number (New) | 8733 | 2454 | 851 | 1161 | 1426 | 203456 |
|  | Peak Number ( Cumulative) | 74605 | 33573 | 5186 | 9787 | 8014 | 5687563 |
|  | End Time | 6/7/2020 | 6/3/2020 | 6/3/2020 | 6/3/2020 | 6/3/2020 | 6/7/2020 |
|  | End Cases | 329467 | 58472 | 12117 | 17285 | 23737 | 6843077 |
|  | Duration | 128 | 120 | 131 | 129 | 131 | 135 |
| Scenario 3 | Peak Time | 4/10/2020 | 4/15/2020 | 3/3/2020 | 4/17/2020 | 4/20/2020 | 5/18/2020 |
|  | Peak Number (New) | 8733 | 2454 | 851 | 1161 | 1426 | 202443 |
|  | Peak Number ( Cumulative) | 74605 | 33573 | 5186 | 9787 | 8014 | 5686550 |
|  | End Time | 7/1/2020 | 7/1/2020 | 7/1/2020 | 7/1/2020 | 7/1/2020 | 7/1/2020 |
|  | End Cases | 447797 | 73541 | 14747 | 21194 | 29297 | 10007832 |
|  | Duration | 152 | 148 | 159 | 157 | 159 | 159 |

## Conclusion/Discussion

As an alternative to the epidemiologic transmission models, we formulated the real-time forecasting and evaluating multiple public health intervention problem into a novel causal inference problem. We viewed the interventions as treatments where multiple interventions were administered at different time points. The number of new cases were taken as treatment responses. The RIN uses sequence-to-sequence multi-input/output recurrent neural network as a tool for modeling the real-time trajectory of the transmission dynamics of Covid-19, health intervention planning and making multi-step prediction of the response trajectory of Covid-19 over time with multiple interventions. The RNN can learn the complex dynamics within the temporal ordering of input time series of Covid-19 and use an internal memory to remember the hidden features.

This AI and causal inference-inspired approach allows us to address three important questions. The first question is the prediction accuracy. Unlike other dynamic systems where the parameters in the systems and control variables are, in general, independent, the epidemic systems have intervention and system dependent parameters. We designed the intervention variable that quantified comprehensive intervention strategies and had close relationships with the parameters in the epidemic systems. Therefore, the RINs could take the parameters in the epidemic dynamic systems as input control variables that can be estimated in the RIN training. The RIN models were closer to real epidemic dynamic systems than the epidemiological models. Therefore, our results showed that the RIN substantially improved the accuracies of prediction and subsequently multiple-step forecasting.

The second question is how important is the intervention time. Since interventions are complicated and are difficult to quantify, we designed three intervention scenarios to represent the degrees and delays of interventions. Since the proposed methods combine the real data and models, they allowed us to evaluate the consequences of multiple intervention strategies, while maintaining the analysis as close to the real data as possible. The RIN investigated the impact of multiple public intervention plans and intervention measures on the size, duration and time of the virus outbreak and recommended the appropriate intervention times.

We estimated the duration, peak time and ending time, peak number of new cases and cumulative cases, and maximum number of cumulative cases of COVID-19 under three intervention scenarios for 184 countries in the world. We observed that the number of cumulative cases of Covid-19 would exponentially grow and reach extremely high number 199,554,596 on July 6, 2020 if none of additional comprehensive public health intervention was implemented. However, we also found that top 12 countries with the largest number of the lab confirmed cumulative cases of COVID-19 were closing to stopping the spread of Covid-19 if additional interventions such as wearing face masks were implemented. We can see that under all three scenarios, the peak times were before April 15, 2020 and the spread of COVID-19 would be sopped before the end of May, 2020.

## Data Availability

Data on the number of cumulative and new cases and COVID-19-attributed deaths across 184 countries from January 22, 2020 to April 7 were obtained from John Hopkins Coronavirus Resource Center (https://coronavirus.jhu.edu/MAP.HTML).

## Acknowledgements

Dr. Li Jin was partially supported by National Natural Science Foundation of China (91846302) Dr. Wei Lin is supported by the National Key R&D Program of China (Grant no. 2018YFC0116600), by the National Natural Science Foundation of China (Grant no. 11925103) and by the STCSM (Grant no. 18DZ1201000).

Thank the reviewers for their helpful comments that substantially improve the paper.

## Supplementary Materials

### Supplementary Note A

#### Recurrent Intervention Network (RIN)

##### Simple RNN Unit

RIN consisted of an encoder RNN and a decoder RNN. The RNN is a mapping from a sequence space (or time series) to another sequence space (or time series space) and the current output depends on both current input and the whole observation history (whole time series). The RNN extracts short-term local dependency patterns among variables and to discover long-term patterns for time series trends (Lai et al. 2017). However, the traditional autoregressive methods do not distinguish the two types of patterns and capture their interactions explicitly and dynamically.

Basic unit of RNN in both encoder and decoder had three layers: input, recurrent hidden and output layers (Figure S1). The input layer consisted of three types of variables: covariates 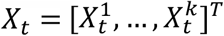, a scaler intervention variable A_*t*_ and the numbers of cases (potential outcomes) *Y*_*t*_ at the time *t*. The covariates can include the rate of virus test, Google mobility indexes. Define the input vector *V*_*t*_ as

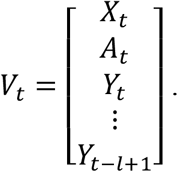

Let 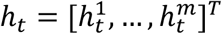 be a *m* dimensional hidden state vector where *m* is set to be 100 in this study. The data *V*_*t*_ is inputted into the input layer. The linear transformation *W*_*vh*_*V*_*t*_ of the data *V*_*t*_ is then sent to the hidden layer, where *W*_*vh*_ is a *m* × (*k* + *l*+ 1) dimensional matrix. The hidden layer receives information from the input layer and hidden layer at the previous time point. The state is determined by the following nonlinear transformation of its received information:

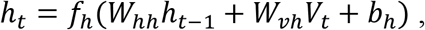

where *W*_*hh*_ is a *m* × *m* dimensional weight matrix that connect the previous state to the current state, and 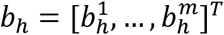 is a *m* dimensional bias vector that corrects the bias, and *f*_*h*_ is a element-wise nonlinear activation function and is often defined as the following “tanh” function:

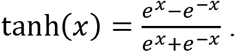

The neurons in hidden layer are connected to the output layer via a *m* dimensional weight vector *W*_*hy*_. The output *Ŷ*_*t*+1_ is determined by

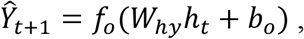

where *f*_*o*_ is an activation function and *b*_*o*_ is the bias vector of the output neurons.

#### Long Short Term Memory Network Unit

Long Short Term Memory network (LSTM) is a special kind of RNN, capable of learning long-term dependencies (Hochreiter and Schmidhuber, 1997; Srivastava et al. 2015). The LSTM attempts to preserve long term useful information and skip short-term irrelevant information. Each LSTM unit has a cell that is a memory unit (Figure S2). The key to LSTMs is the cell state. The cell state is determined by four gates: forget gate, input gate, input modulation, and output gate. At each time step, the unit receives input from two external sources: the current input variables 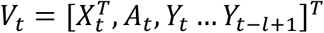 and the previous hidden states *h*_*t*−1_. The total input activates the gates through sigmoid function and the tanh nonlinear function. The forget gate layer decides what information we’re going to throw away from the cell state:

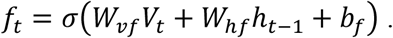

The next step, the input gate layer decides which values we’ll update:

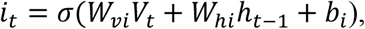

Next, input modulation gate creates a vector of new candidate values, *g*_*t*_, that could be added to the state:

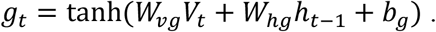

The old cell state is updated by

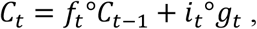

where ° denotes element-wise multiplication.

Output gate decides what parts of the cell state we’re going to output:

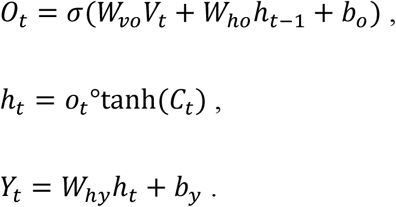

We update the hidden state *h*_*t*_ by tanh transformation of cell state, multiplied by the output of the sigmoid output gate. When the output gate is 1, we pass all memory information in the cell state through to the predictor. In contrast, when the output is 0, we keep all the information only within the cell.

#### Potential Outcome Framework for Evaluation of Public Health Interventions

Learning intervention policies is an extremely challenge problem. We employ the Counterfactually-Guided Policy Evaluation (CF-GPS) principle to evaluate the effect of public health interventions on controlling the spread of Covid-19 (Buesing et al. 2018). We view the public health intervention as treatment and the number of cases as the outcome. Counterfactual treatment outcome estimation is essentially a causal problem. Most methods for causal inference are designed for the statistic setting and cannot be applied to evaluating the effects of the sequence of public health interventions on (e.g. sequential application of intervention A followed by intervention B) the transmission dynamics of Covid-19 over time. The models that can deal with varying-length transmission dynamic histories of Covid-19 are needed for estimating intervention effects over time.

Potential outcome framework is our basic model to evaluate the impact of the public health interventions on the spread of Covid-19. The potential outcome framework is often referred to the Neyman-Rubin model (Rubin 1974). Potential outcomes consist of actual (or observed) and counterfactual (hypothesized) outcomes. We are interested in number of cases of Covid-19 under some specific intervention. We observed the number of cases of Covid-19 (actual observation) without intervention or known intervention. However, we want to know what number of cases of Covid-19 (counterfactual, unobserved) would be if an alternative intervention was implemented. To evaluate the effect of intervention, we should compare the difference between the observed actual number of cases of Covid-19 and the counterfactual number of cases of Covid-19. Our aim is to learn the counterfactual outcomes of Coid-19 under a sequence of public health intervention options and evaluate the impact of the intervention strategies on the spread of Covid-19.

The Recurrent Intervention Network (RIN), a novel sequence-to-sequence recurrent neural network is an architecture for estimating the effects of intervention on the spread of Covid-19 over time. A basic block of the RIN is the RNN. The RNN avoids the need for specifying any explicit model, using the networks to learn the relationships among variables directly from the data. The RIN consists of an encoder and a decoder. Both of the encoder and decoder use the RNN as their basic architectures (Bica et al. 2020).

The encoder network uses vanilla RNN, or LSTM to model the history of the covariates, interventions and outcomes (number of cases), learn the causal structure from the retrospective data and predict one-step-ahead outcomes, given observations of covariates and actual interventions.

The decoder network uses the hidden state computed by the encoder to initialize the state of an RNN in the decoder which predicts the counterfactual outcomes for a sequence of hypothesized future interventions (Bica et al. 2020). The decoder attempts to propagate the encoder representation forwards in time, using only the planned interventions and avoiding the covariates.

#### Problem Mathematical Formulation

Consider *N* countries in the world. Each country is viewed as a sample. For the *i*^*th*^ country and time point *t*, let 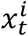 be a set of covariates and 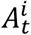 be an intervention. The intervention variable 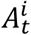 can be a binary variable: 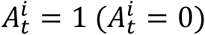 indicates that intervention is (not) implemented. The intervention variable 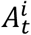 can also be continuous variable taking values in the interval [0, 1]. If 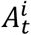 is a continuous variable, the value of 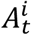 represents the intensity of intervention. 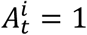 indicates that the intervention is the most strict and comprehensive public health intervention. 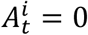 indicates that no intervention is taken. Let 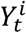 be the outcome (number of cases) of the *i*^*th*^ country at the time point *t*. The observed dataset is denoted by 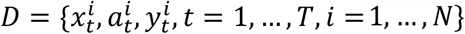 where 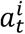 is prespecified.

The potential outcome framework for treatment effect estimation which accounts for the time varying treatments (Bica et al. 2020) is extended to evaluation of the effects of the interventions on the spread of Covid-19. Let 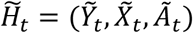 be the history of the outcomes (number of cases) 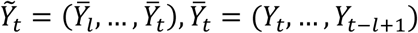, the covariates 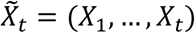 and interventions *Ã*_*t*_ = (*A*_1_, …, *A*_*t*_). Let *Y*[*ã*] be the potential outcomes that can be either observed or counterfactual, under each possible sequence of intervention *ã*. The potential outcome framework assumes the existence of the hypothetical outcome with some interventions which is not observed in the data. The hypothetical outcome under hypothetical intervention is called counterfactual outcome. Given the history 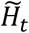 and a sequence of planned interventions *Ã*(*t, t* + *τ* − 1) = (*A*, …, *A*_*t*+*τ*−1_), the counterfactual outcome *Y*_*t*+*τ*_[*Ã*(*t,t* + *τ* − 1)] can be predicted by

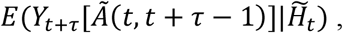

which define the future dynamic trajectory of the Covid-19 under the planned sequence of interventions, given the previous history of Covid-19 and its environments. To make the prediction of the dynamic trajectory of the Covid-19 under the potential outcome framework to be identifiable, we need to make the following assumptions (Bica et al. 2020).

#### Assumptions

We introduce assumptions in the Neyman-Rubin model (Bica et al., 2020).

##### Assumption 1. (Consistency)

If a nation receives an intervention A_*t*_ = *a*_*t*_, then the potential outcome for the intervention *a*_*t*_ which can be counterfactual is equal to the observed (factual) outcome *Y*_*t*+1_(*a*_*t*_) = *Y*_*t*+1_.

##### Assumption 2. (Overlap)

For all (*a*_1_, *x*_1_, …, *a*_*t*−1_, *x*_*t*−1_), we have 0 < *P*(A_*t*_ = *a*_*t*_|A_1_ = *a*_1_, *X*_1_ = *x*_1_, …, A_*t*−1_ = *a*_*t*−1_, *X*_*t*−1_ = *x*_*t*−1_) < 1. In other words, at each time step, each intervention has non-zero probability of being implemented.

##### Assumption 3. Sequential strong ignorability

Conditional on A_1_ = *a*_1_, *X*_1_ = *x*_1_, …, A_*t*−1_ = *a*_*t*−1_, *X*_*t*−1_ = *x*_*t*−1_, the potential outcomes *Y*_*t*+1_ are independent of A_*t*_,

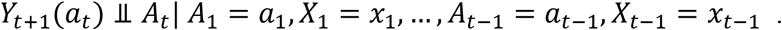

Assumption 3 implies that there is no confounders which affect both outcomes and interventions.

#### Training Procedures and Loss function

As we discussed in the previous section, the counterfactual policy evaluation (CPE) was a powerful tool to address the pressing issue of evaluating the performance of public health interventions. The CPE problem was formulated as a potential outcome or counterfactual estimation problem (Bibaut et al. 2019). The RIN training consisted of the encoder training and decoder training. We first fitted a model of the system’s dynamics of Covid-19 to the data from past experimental interventions to learn representations of the states of the dynamics of Covid-19 (encoder training), and then used the learned fit to extrapolate and forecast the response to the alternative interventions (decoder training).

Since the output is continuous, the mean square errors are used as the loss function for the encoder:

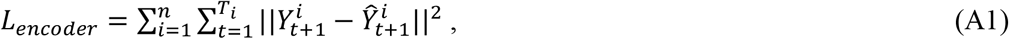

where *n* is the number of countries with outbreak of Covid-19 and *T*_*i*_ is the length of the observed country’s trajectory of Covid-19.

For the decoder, we assumed that observations were batched into shorter sequences of up to *τ*_*b*_, a prediction horizon. The loss function for the decoder is defined as

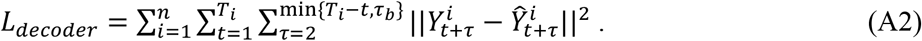

The major steps for encoder and decoder training were given below.

#### Step 1: Data pre-processing

Data were split into training dataset (01/22-04/27, 2020) and validation dataset (04/28-05/08/2020). All the input number of lab-confirmed cumulative cases *Y*_*t*_ was pre-processed by the following transformation: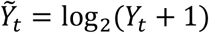. The number of new cases was calculated as 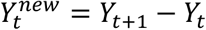.

#### Step 2: Generate the intervention measure curve

The intervention measure was calculated as follows. Set the intervention measure at the final time 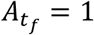 for China, 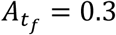 for Korea South, Switzerland, United Kingdom, Spain, US, Italy, Germany, Iran, and France, and 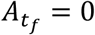 for all other countries. Assume that The intervention measure curve is an exponential function starting at 0 and ends at 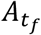 The intervention measure *A*_*t*_ is given by 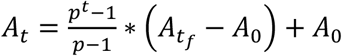, where *p* > 0, *p* ≠ 1 is the curve shape factor and *t* takes values in evenly sliced numbers of interval [0, 1], *A*_0_ is the intervention measure at the initial time *t*_0_. When *p* = 1, *A*_*t*_ is a linear function 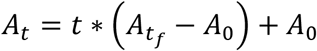. In this study, we set *p* = 0.01.

##### Step 3: minibatches and normalization

Randomly picked *k* = 64 countries with *l*= 7 length of Covid-19 time series data staring from the same day to generate *k* time series with *l* length for a minibatch to be used for backpropagation training through time. Calculate the mean value of each time series in the batch. The values of each time series were divided by their mean values.

#### Step 4: Encoder Training

The randomly grouped minibatch data with *k* = 64 countries and *l* = 7 length of the number of lab confirmed new cases of Covid-19 and sequence of interventions were used as input to train the encoder. The standard back-propagation method and the Adam Optimizer were used to minimize the loss function of each batch, defined in equation (A1). Upon completion, the encoder was used to perform a feed-forward pass over the training data for extraction of the internal states *h*_*t*_ that were used to train the decoder and for prediction of one-step-ahead outcomes *Y*_*t*+1_.

#### Step 5: Decoder Training

During decoder training, the decoder uses as input the extracted internal states *h*_*t*_ computed by the encoder, the outcomes (*Y*_*t*+2_, …, *Y*_*t*+*τ*_) from the observational data that were batched into a shorter sequences of up to *τ* steps, and the planned sequence of interventions (*A*_*t*+1_, …, *A*_*t*+*τ*−1_). Similar to encoder training, the standard back-propagation method and the Adam Optimizer were used to minimize the loss function of each batch, defined in equation (A2). The initial learning rate in the updating parameters in both encoder and decoder training was 0.02 and learning rate decay was 0.0001.

#### Step 6: Evaluation of the Intervention Strategies and Forecasting

After completion of the training, the decoder can be used to evaluation of interventions. During evaluation, we do not have access to ground-truth outcomes. Therefore, we used the decoder to make one step ahead forecasting. The outcomes forecasted by the decoder (*Ŷ*_*t*+1_, …, *Ŷ*_*t*+*τ*−1_) were recursively used as inputs. For each country, by running the decoder that was trained in step 5, with a sequence of planned interventions and recursively forecasted outcomes, we forecasted the response dynamics, i.e., the number of new cases of Covid-19 of the country over time under a sequence of interventions and evaluate the effects of various intervention strategies on controlling the spread of Covid-19. By running the decoder, we can select starting and ending time of different interventions and the optimal or appropriate interventions to give over time to obtain the best outcomes of controlling the spread of Covid-19 for each country.

---

**Figure S1.**
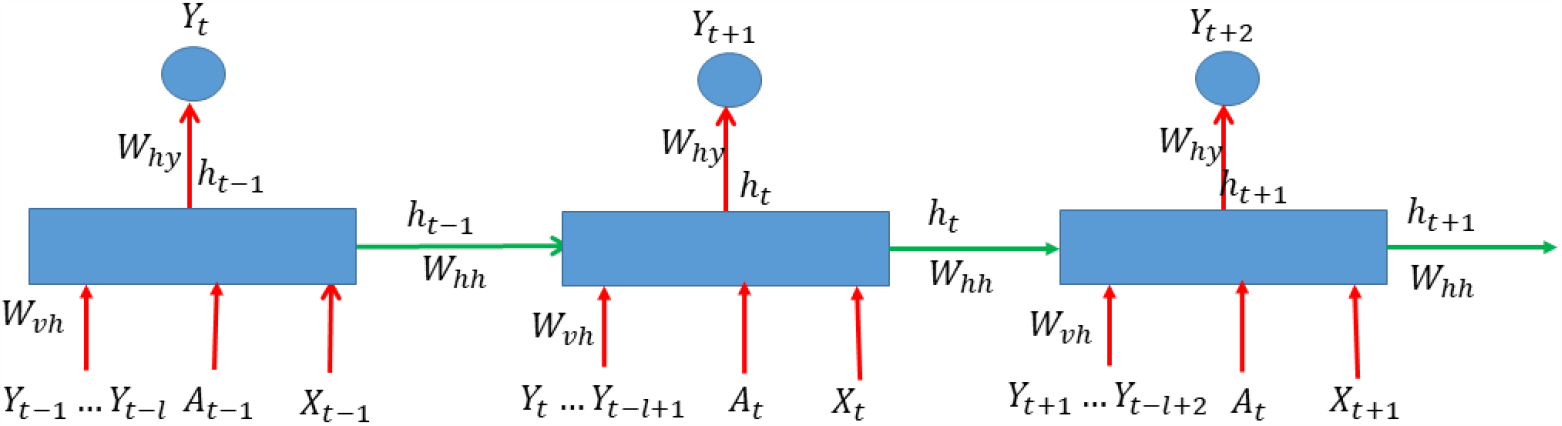
Architecture of vanilla RNN.

**Figure S2.**
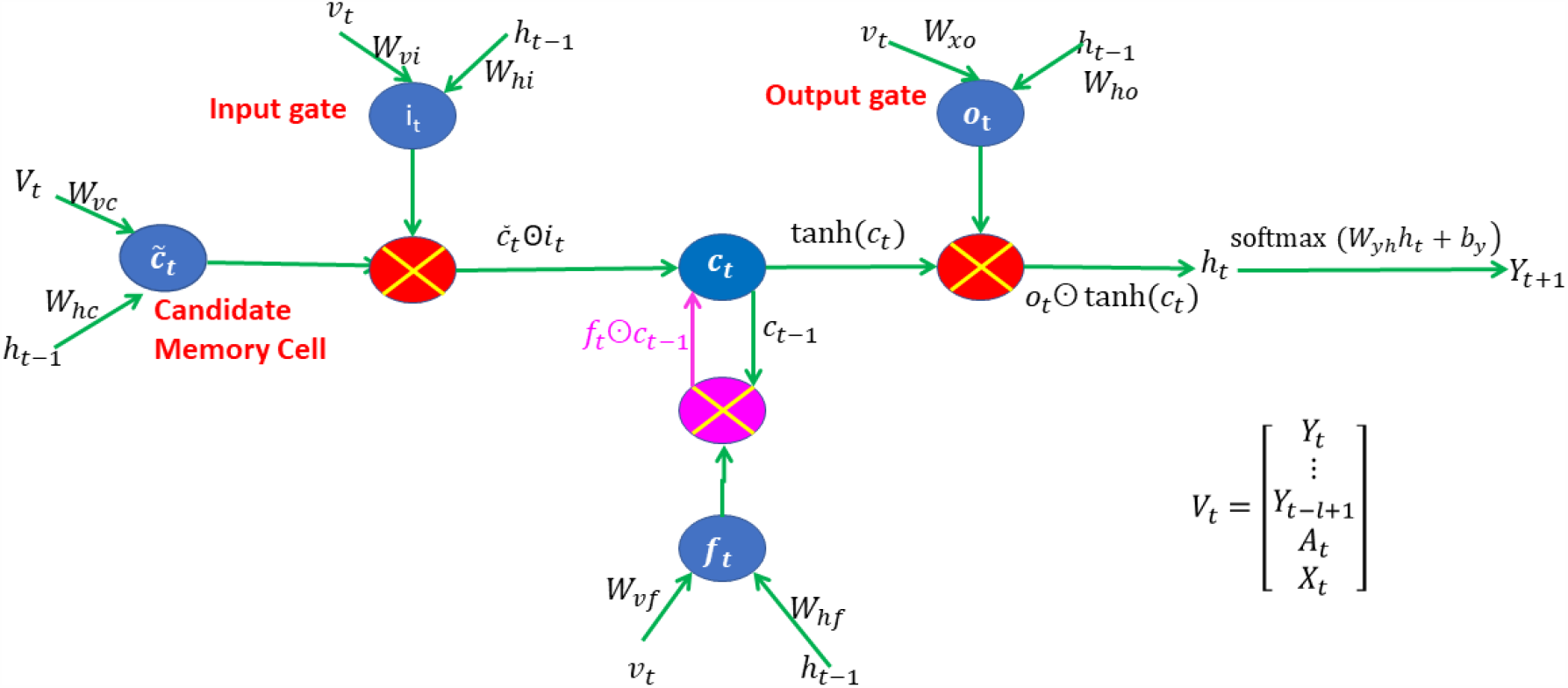
Architecture of LSTM.

**Figures S3.**
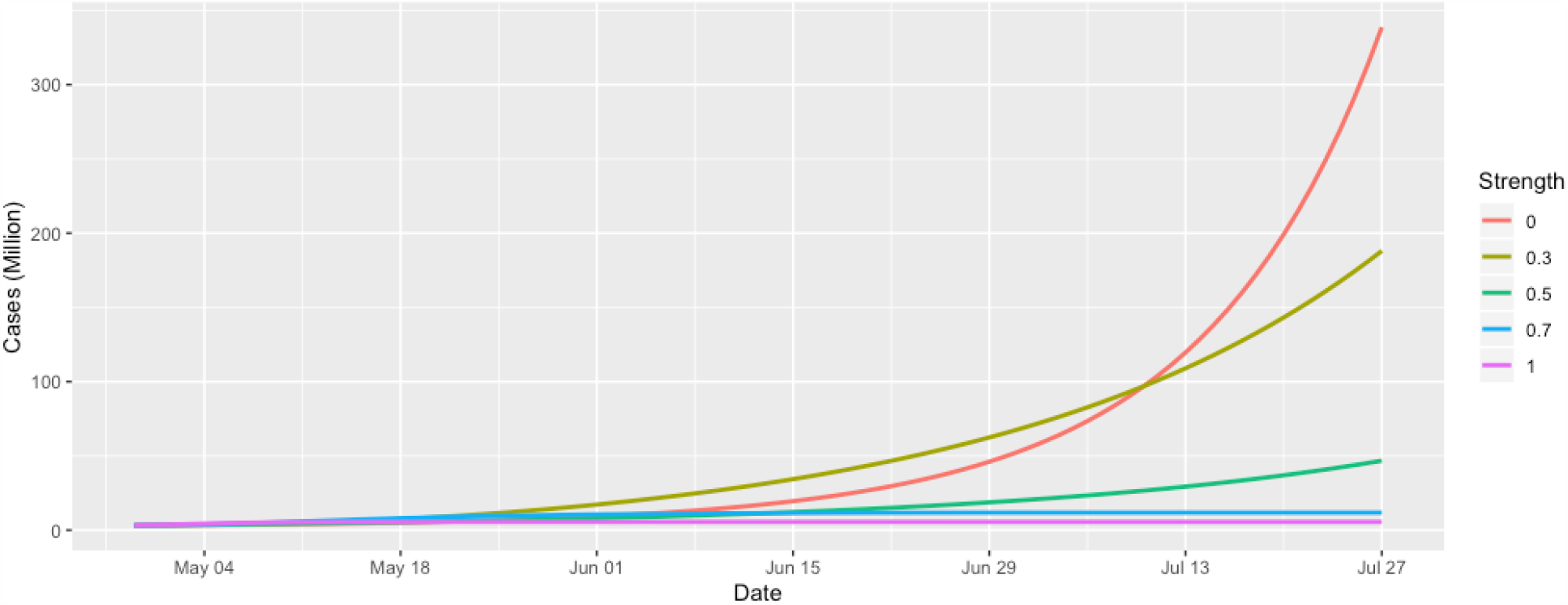
Counterfactual numbers of cases of Covid-19 over time worldwide to respond the interventions with values 0, 0.3, 0.5, 0.7 and 1.

**Figure S4.**
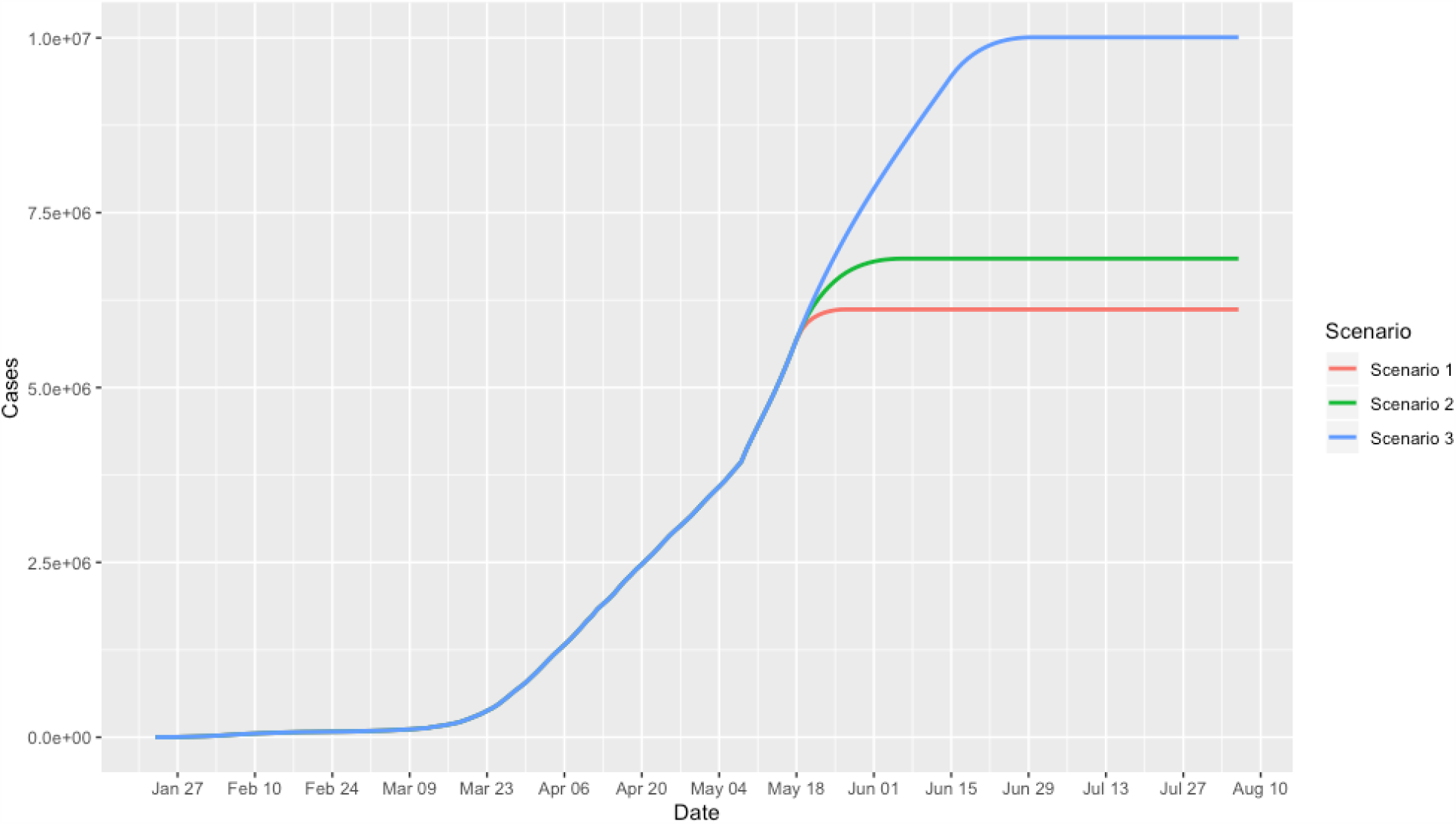
Time-case plot of Covid-19 worldwide under three invention scenarios

